# Drivers of geographic patterns in outpatient antibiotic prescribing in the United States

**DOI:** 10.1101/2023.10.25.23297553

**Authors:** Stephen M. Kissler, Kirstin I. Oliveira Roster, Rachel Petherbridge, Ateev Mehrotra, Michael L. Barnett, Yonatan H. Grad

## Abstract

Antibiotic prescribing rates vary dramatically across regions in the United States, but the relative importance in prescribing practices versus per capita visit rates in driving this variation remain unclear. Quantifying their relative importance can aid in reducing antibiotic use. Using US medical claims data from 2015-2018 covering over 15 million privately insured adults, we conducted a regression analysis to estimate the relative contribution of outpatient visit rates and per-visit prescribing in explaining variation in outpatient antibiotic prescribing rates across metropolitan statistical areas. Variation in visit rates per capita explained more of the geographic variation in outpatient antibiotic prescribing rates than per-visit prescribing for conditions with high prescribing volume. Efforts to reduce antibiotic use may benefit from addressing the factors driving higher rates of outpatient visits, in addition to continued focus on stewardship.

## Introduction

Reducing antibiotic consumption is a major public health objective. Various interventions have been deployed to ensure that physicians only prescribe antibiotics for appropriate conditions (1,2), yet there remains substantial antibiotic use (3,4). Reducing antibiotic consumption may require looking beyond clinical decisions about a patient’s need for antibiotics to the upstream factors that influence the rates of medical encounters that lead to antibiotic prescriptions, such as the prevalence of infectious diseases and access to care.

Geographic comparisons can clarify the relationship between these upstream factors and antibiotic prescribing. In the United States, outpatient antibiotic prescribing differs substantially across regions, with the highest rates in the South and the lowest in the West (5,6). This tracks with the prevalence of some infections, such as gonorrhea (7) and invasive pneumococcal disease (8), raising the possibility that differences in antibiotic prescribing rates may be in part due to differences in disease burden.

Prior work has found that outpatient visit rates in Massachusetts were closely linked with both geographic (9) and temporal (10) variation in antibiotic prescribing. However, it is not clear whether the same pattern holds nationally. Using a large medical insurance claims database, we examined the association between antibiotic prescriptions per capita and outpatient visit rates versus rates of per-visit prescribing.

## Methods

### Study design

Data on outpatient visits and pharmacy claims for antibiotic prescriptions were extracted from the Merative™ MarketScan® Commercial Database for the period from 2015 to 2018. These data include health insurance claims from outpatient care and pharmacy data as well as enrollment data from large employers and health plans across the United States who provide private healthcare coverage for 15-19 million employees, their spouses, and dependents each year (11). Regional, sex, and age distributions in the data were comparable to the whole population (12) (**Supplementary Table S1**).

For each person, we extracted all available outpatient and pharmacy claims. Outpatient claims included up to two diagnoses coded according to the International Classification of Diseases (ICD). We aggregated these diagnoses into broader classes (“conditions”) using mappings provided by the Agency for Healthcare Research and Quality’s (AHRQ’s) Clinical Classification Software (CCS) (13). Pharmacy claims included a National Drug Code (NDC) identifier. We used these identifiers to extract all new (*i.e*., non-refill) antibiotic prescriptions (**Supplementary Methods**).

The MarketScan database does not explicitly link pharmacy claims with the medical conditions that prompted them. To address this, we linked each claim with the most recent outpatient visit that occurred within the prior seven days (**Supplementary Methods**) (9). Prescriptions without outpatient visits in the seven days prior were labelled as “unlinked” and excluded from the analysis. For visits with multiple diagnoses, we chose the medical condition that was most likely to have prompted the antibiotic prescription, following prioritization schemes used in previous studies (9,14) (**Supplementary Table 2**).

### Study outcomes

We estimated (1) the annual number of antibiotic prescriptions per 1,000 individuals, overall, by region, and by metropolitan statistical area (MSA) and (2) the degree to which outpatient visit rates per 1,000 individuals and per-visit prescribing rates each explain geographic variation in antibiotic prescription rates.

### Statistical analysis

We calculated the number of antibiotic prescriptions per person-year, the number of outpatient visits per person-year, and the number of antibiotic prescriptions filled per visit, stratified by medical condition and MSA. To assess the relative contribution of outpatient visit rates and per-visit prescribing to the number of antibiotic prescriptions per person-year, we calculated the coefficients of determination (*R*^*2*^) for individual regressions of antibiotic prescriptions per person-year for each condition on (a) outpatient visits per person-year and (b) antibiotic prescriptions per visit, with higher *R*^*2*^ indicating a greater amount of variation explained. We focused on the five conditions with the highest prescribing volume in the main analysis and conducted a sensitivity analysis of all conditions (**Supplementary Figures 1 and 2**). Analyses were conducted using R version 4.1.2 and Python version 3.9.13.

**Figure 1.**
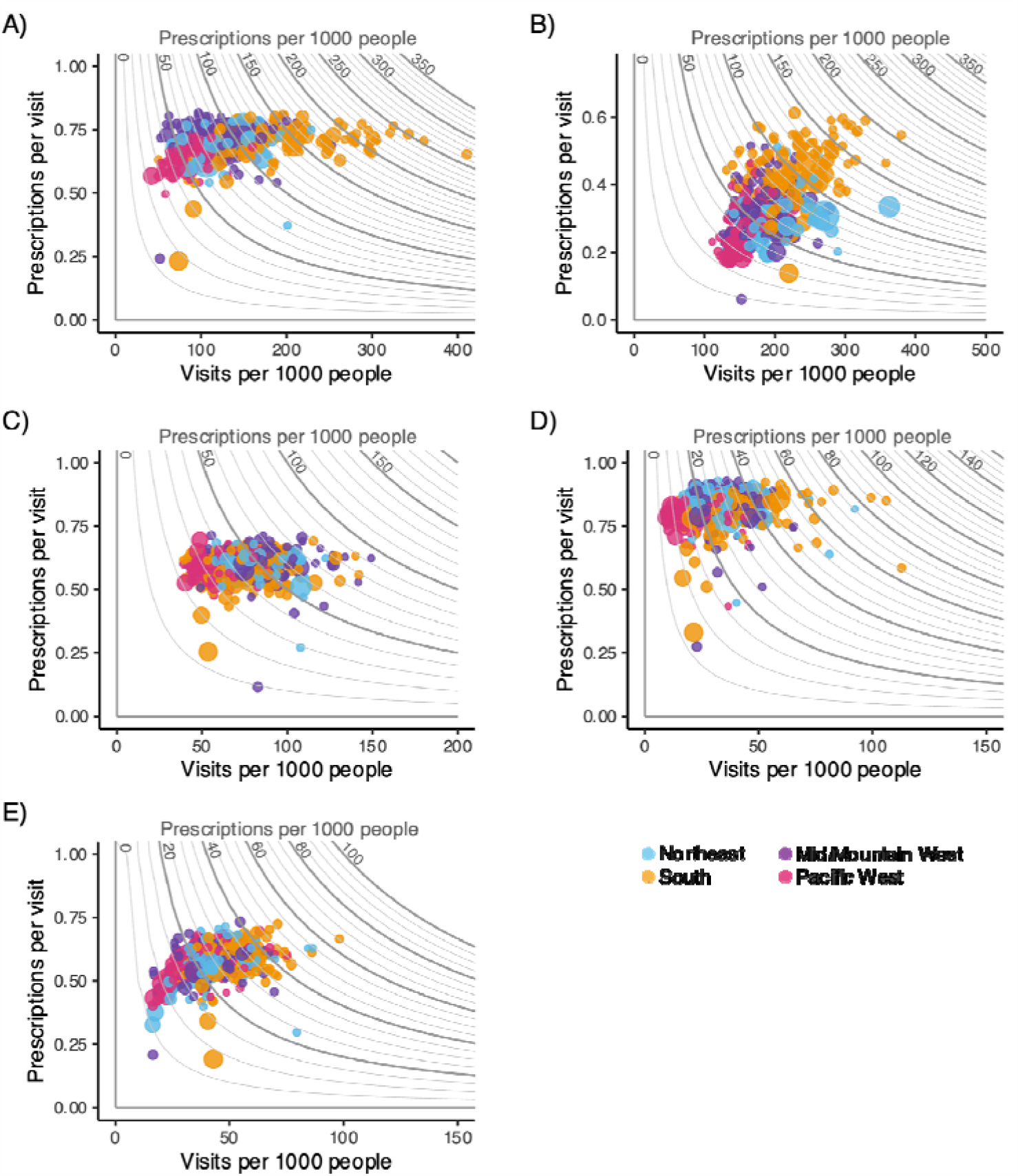
Variation in outpatient visits, per-visit prescribing, and *per capita* antibiotic prescribing rates across the US. (A-E) Contour plots depicting outpatient visit rate per 1,000 individuals (horizontal axis), per-visit prescribing rate (vertical axis), and antibiotic prescribing rate per 1,000 individuals (contours, equal to the product of the outpatient visit rate and the per-visit prescribing rate) for MSAs (points) in the United States. Points are colored according to the region where the MSA is geographically situated. Point sizes are proportional to the number of individuals from that MSA in the study population. Contour plots for (A) Sinusitis, (B) other URI, (C) otitis media, (D) streptococcal pharyngitis, and (E) acute bronchitis are depicted here. Contour plots for the remaining conditions are included in the **Supplementary Material**.

### Data and code availability

Full data and code are available at: https://github.com/gradlab/AbxGeography

## Results

The five conditions with the highest prescribing volume—sinusitis, otitis media, streptococcal pharyngitis, acute bronchitis, and other upper respiratory tract infections (URIs)—accounted for a total annual mean of 263.9 antibiotic prescriptions per 1,000 people, comprising 52.5% of all new antibiotic prescriptions filled between 2015 and 2018 (**Table 1**).

**Table 1.** Antibiotic prescribing rates associated with the five most highly prescribed medical conditions, overall and by US region. Darker shading indicates greater values, with each variable represented by a different color to account for differences in units.

| Condition | Antibiotic prescriptions per 1,000 people | Percent of total antibiotic prescriptions | Visits per 1,000 people | Percent of visits with an antibiotic prescription |
| --- | --- | --- | --- | --- |
| Sinusitis | 97.1 | 19.3 | 139.0 | 69.9 |
| Northeast | 81 | 2.8 | 120.8 | 67.1 |
| South | 120.3 | 10.3 | 170.5 | 70.5 |
| Midwest | 92.1 | 4.1 | 127.5 | 72.2 |
| West | 61.9 | 2.1 | 93.4 | 66.2 |
| URI (other) | 71.5 | 14.2 | 208.1 | 34.4 |
| Northeast | 69.1 | 2.4 | 235.3 | 29.4 |
| South | 91.1 | 7.8 | 227.9 | 40.0 |
| Midwest | 57.0 | 2.5 | 184.4 | 30.9 |
| West | 43.8 | 1.5 | 162.5 | 27.0 |
| Otitis media | 44.7 | 8.9 | 75.8 | 59.0 |
| Northeast | 44.8 | 1.5 | 77.8 | 57.6 |
| South | 47.3 | 4.1 | 80.6 | 58.7 |
| Midwest | 48.3 | 2.1 | 80.1 | 60.3 |
| West | 33.6 | 1.2 | 56.2 | 59.8 |
| Strep pharyngitis | 26.6 | 5.3 | 32.2 | 82.5 |
| Northeast | 26.1 | 0.9 | 31.5 | 82.8 |
| South | 32.0 | 2.8 | 38.9 | 82.3 |
| Midwest | 25.1 | 1.1 | 30.0 | 83.7 |
| West | 15.3 | 0.5 | 19.0 | 80.7 |
| Bronchitis (acute) | 24.0 | 4.8 | 41.8 | 57.5 |
| Northeast | 21.9 | 0.8 | 38.8 | 56.4 |
| South | 28.0 | 2.4 | 47.9 | 58.4 |
| Midwest | 23.5 | 1.0 | 40.6 | 57.8 |
| West | 17.1 | 0.6 | 31.0 | 55.1 |

Antibiotic receipt per 1000 people was highest in the South for each of these conditions except otitis media, which was highest in the Midwest. Outpatient visits rates associated with these conditions varied substantially across regions and were consistently highest in the South, except for “other URI,” which were highest in the Northeast (**Table 1**). For example, visits *per capita* for Streptococcal pharyngitis were more than double in the South compared to the West (38.9 and 19 visits per 1000 persons, respectively; **Table 1**). Per-visit prescribing varied less among regions, and there was no region with consistently higher per-visit prescribing (**Table 1**).

For the five conditions with the highest prescribing volume, outpatient visit rates accounted for a higher degree of the geographic variation in prescribing rates than per-visit prescribing for those conditions (**Figure 1**). For sinusitis, for example, (**Figure 1A**), *per capita* outpatient visits explained a high degree of variation in sinusitis-related antibiotic prescribing rates across MSAs (*R*^*2*^ = 0.96), while per-visit prescribing rates explained relatively less variation (*R*^*2*^ = 0.16, **Supplementary Figure 2**). Similar patterns were observed for otitis media, streptococcal pharyngitis, and acute bronchitis. For “other URI” (**Figure 1B**), rates of *per capita* outpatient visits and per-visit prescribing described a similar degree of variation in prescribing rates across MSAs (*R*^*2*^ = 0.77 and 0.71, respectively; **Supplementary Figure 2**). For intestinal infections and other conditions for which antibiotics were less frequently prescribed, per-visit-prescribing explained more of the variation in antibiotic prescribing rates than did visit rates (**Supplementary Figure 2**).

## Discussion

Regional differences in antibiotic receipt per capita are driven by the combination of differences in rates of visits and differences in rates of per-visit prescribing. For the most common conditions that result in antibiotic prescribing, geographic variation in outpatient visit rates explained most of the variation in antibiotics receipt per capita.

Prior research has been based on the implicit assumption that regional variation in antibiotic prescribing rates is attributable to differing rates of over-prescribing (14). Our results challenge that assumption. Separating prescribing rates into visit rates (a measure combining disease prevalence and care seeking behavior) and per-visit prescribing rates (a measure of stewardship) showed that per-visit prescribing does not account for much of the geographic variation in prescription fills for the conditions that account for the majority of antibiotic prescriptions. It is unclear what drives the variation in visit rates. Factors could include socioeconomic status, disease prevalence, vaccination, societal expectations on when one should receive care, and access to care. Future research should explore these factors in more depth, but such analyses are currently limited by a lack of data that capture these factors together with visits and prescribing at a fine geographic resolution. Investments in improved data collection and data linkage will be necessary to develop effective intervention targets to reduce the risks of antibiotic resistance.

These results might point to interventions to reduce antibiotic prescribing. For the most common conditions that result in antibiotic prescriptions, interventions may need to target the need for visits to reduce antibiotic use, such as vaccination (15) or reducing delays in care among underserved populations (16).

## Data Availability

Full data and code are available at: https://github.com/gradlab/AbxGeography.

## Funding statement

S.M.K received funding from NIH T32 training grant 2 T32 AI 7535-21 A1. K. I. Oliveira Roster gratefully acknowledges the support of the São Paulo Research Foundation (FAPESP) under grant 2021/11608-6. R.P. received funding from NIH T32 training grant T32 GM135014. This project has been funded in part by contract 200-2016-91779 from the US Centers for Disease Control and Prevention to YHG. Disclaimer: The findings, conclusions, and views expressed are those of the author(s) and do not necessarily represent the official position of the Centers for Disease Control and Prevention (CDC).

## Conflicts of Interest

The authors declare no conflicts of interest.

## Data availability

Full data and code for this analysis are available at https://github.com/gradlab/AbxGeography

## Notes

### Competing Interest Statement

The authors have declared no competing interest.

