## supplement for "Drivers of geographic patterns in outpatient antibiotic prescribing in the United States"

### Supplementary Materials.

#### Supplementary Methods.

Linking pharmacy claims with outpatient visits. Outpatient claims included up to two diagnoses coded according to the International Classification of Diseases (ICD). We aggregated these diagnoses into broader classes ("conditions") using mappings provided by the Agency for Healthcare Research and Quality's (AHRQ's) Clinical Classification Software (CCS)  To identify individual outpatient visits, we extracted the first outpatient encounter associated with each person on a given day. Frequenly, there are many claims on a given day associated with a single encounter, capturing various procedures that are all associated with a common diagnosis or set of diagnoses. Then, for each person, we extracted all pharmacy claims and restricted to antibiotics on the basis of National Drug Code (NDC) codes. For each antibiotic claim, we found the most recent outpatient visit that occurred for that person within the preceding seven days (including the day of prescription fill). This constituted a "link" between the prescription and an outpatient visit. If there was no outpatient visit in the seven days preceding the prescription fill, the prescription was marked as "unlinked", affecting less than 15% of prescriptions. Mappings from ICD diagnoses to CCS conditions and from NDC codes to antibiotics are included in the online code repository associated with this manuscript: https://github.com/gradlab/AbxGeography.

Sensitivity analysis of all conditions. As in the main analysis, we also measured the relative contribution of outpatient visit rates and per-visit prescribing to the geographic variation in antibiotic prescribing rates, by calculating the coefficient of determination ($R^2$) for the regression of antibiotic prescriptions per person-year on (a) outpatient visits per person-year and (b) antibiotic prescriptions per visit, with higher $R^2$ indicating a greater amount of variation explained (Supplementary Figure S1). Additionally, we measured how the amount of variation explained by outpatient visit rates and per-visit prescribing rates was related to the total prescribing volume across all conditions in the dataset. Specifically, we conducted an additional regression of the $R^2$ values for each of the two variables against the $\log_{10}$–transformed total volume of antibiotic prescriptions associated with each condition, to assess whether the explanatory power of outpatient visits and per-visit prescribing rates varied systematically by the total volume of antibiotics prescribed for a given condition (Supplementary Figure S2).

**Supplementary Table 1. Characteristics of the study population and reference values from the 2020 Census**

| Variable | Category | Number of people (%) by year | | | | Reference values |
|---|---|---|---|---|---|---|
| | | 2015 | 2016 | 2017 | 2018 | 2020 Census |
| Total | – | 18989691 (100%) | 18897347 (100%) | 15574072 (100%) | 16419151 (100%) | |
| Sex | Male | 9139343 (48.1%) | 9113395 (48.2%) | 7594325 (48.8%) | 8061316 (49.1%) | 49.1% |
| | Female | 9850348 (51.9%) | 9783952 (51.8%) | 7979747 (51.2%) | 8357835 (50.9%) | 50.9% |
| Census Region | Northeast | 3401805 (17.9%) | 3071460 (16.3%) | 2351782 (15.1%) | 3173820 (19.3%) | 17.2% |
| | South | 8393206 (44.2%) | 8679832 (45.9%) | 6671373 (42.8%) | 6425726 (39.1%) | 38.4% |
| | Midwest | 3916502 (20.6%) | 3862332 (20.4%) | 3863799 (24.8%) | 3965199 (24.1%) | 20.7% |
| | West | 3278178 (17.3%) | 3283723 (17.4%) | 2687118 (17.3%) | 2854406 (17.4%) | 23.7% |
| Age group (years) | 0-4 | 1018741 (5.4%) | 1023975 (5.4%) | 866737 (5.6%) | 902236 (5.5%) | 6.0% |
| | 5-9 | 1244135 (6.6%) | 1217948 (6.4%) | 1008299 (6.5%) | 1041415 (6.3%) | 6.2% |
| | 10-14 | 1413095 (7.4%) | 1392233 (7.4%) | 1164661 (7.5%) | 1215861 (7.4%) | 6.4% |
| | 15-19 | 1526902 (8.0%) | 1376856 (7.3%) | 1143650 (7.3%) | 1191431 (7.3%) | 6.4% |
| | 20-24 | 1607390 (8.5%) | 1589339 (8.4%) | 1315453 (8.4%) | 1385926 (8.4%) | 6.5% |
| | 25-29 | 1037819 (5.5%) | 1107728 (5.9%) | 937160 (6.0%) | 995527 (6.1%) | 7.1% |
| | 30-34 | 1316622 (6.9%) | 1313278 (6.9%) | 1108050 (7.1%) | 1179238 (7.2%) | 6.8% |
| | 35-39 | 1419642 (7.5%) | 1436661 (7.6%) | 1212464 (7.8%) | 1298883 (7.9%) | 6.6% |
| | 40-44 | 1588943 (8.4%) | 1508533 (8.0%) | 1245861 (8.0%) | 1309912 (8.0%) | 6.1% |
| | 45-49 | 1673758 (8.8%) | 1683088 (8.9%) | 1394427 (9.0%) | 1464421 (8.9%) | 6.0% |
| | 50-54 | 1876276 (9.9%) | 1819797 (9.6%) | 1456097 (9.3%) | 1522409 (9.3%) | 6.1% |
| | 55-59 | 1880816 (9.9%) | 1883292 (10%) | 1509362 (9.7%) | 1605771 (9.8%) | 6.5% |
| | 60-64 | 1385552 (7.3%) | 1544619 (8.2%) | 1211851 (7.8%) | 1306121 (8.0%) | 6.5% |

**Supplementary Table 2. Prioritization scheme for assigning antibiotic prescriptions to outpatient visits.** If a person had multiple diagnoses associated with the most recent visit preceding an antibiotic prescription, the condition with the highest rank in this table was marked as the condition associated with that prescription. For streptococcal pharyngitis and sinusitits (in CCD9), there is no corresponding CCS condition, so these were extracted explicitly using ICD codes. Asterisks denote a wildcard symbol, so that ICD code 461* contains all ICD codes that begin with 461 and end with a single additional number.

| Condition | CCS9 code | CCS10 code |
|---|---|---|
| Pneumonia | 122 | RSP002 |
| UTI | 159 | GEN004 |
| Intestinal infection | 135 | DIG001 |
| Strep pharyngitis | ICD= 4610 | ICD= J020 |
| Sinusitis | ICD=461*, 473* | RSP001 |
| URI (other) | 126 | RSP006 |
| SSTI | 197 | SKN001 |
| Otitis media | 92 | EAR001 |
| Asthma | 128 | RSP009 |
| Bacterial infection (other) | 3 | INF003 |
| Tonsillitis | 124 | RSP004 |
| Bronchitis (acute) | 125 | RSP005 |
| Influenza | 123 | RSP003 |
| Viral infection | 7 | INF008 |
| Essential hypertension | 98 | CIR007 |

**Supplementary Table 3. Antibiotic prescribing rates associated with various medical conditions.**

| Condition | Antibiotic prescriptions per 1,000 people | Percent of antibiotic prescriptions nationally | Visits per 1,000 people | Percent of visits with an antibiotic prescription |
|---|---|---|---|---|
| Sinusitis | 97.1 | 19.3 | 139.0 | 69.9 |
| URI (other) | 71.5 | 14.2 | 208.1 | 34.4 |
| Otitis media | 44.7 | 8.9 | 75.8 | 59.0 |
| Strep pharyngitis | 26.6 | 5.3 | 32.2 | 82.5 |
| Bronchitis (acute) | 24.0 | 4.8 | 41.8 | 57.5 |
| SSTI | 19.7 | 3.9 | 54.9 | 35.9 |
| UTI | 17.7 | 3.5 | 58.3 | 30.4 |
| Pneumonia | 7.5 | 1.5 | 17.9 | 42.2 |
| Essential hypertension | 6.9 | 1.4 | 241.4 | 2.9 |
| Asthma | 5.6 | 1.1 | 60.7 | 9.1 |
| Bacterial infection (other) | 5.3 | 1.0 | 16.1 | 33.0 |
| Tonsillitis | 4.7 | 0.9 | 13.1 | 35.8 |
| Viral infection | 3.3 | 0.7 | 85.6 | 3.8 |
| Influenza | 1.4 | 0.3 | 16.5 | 8.3 |
| Intestinal infection | 1.3 | 0.2 | 11.5 | 11.0 |
| Motor vehicle traffic | 0 | 0 | 0.2 | 1.3 |
| Other | 164.7 | 32.8 | 6881.1 | 2.4 |
| **Total** | **502.1** | **100** | **7954.2** | **6.3** |

**Supplementary Figure 1. Variation in outpatient visits, per-visit prescribing, and per capita antibiotic prescribing rates across the US.**

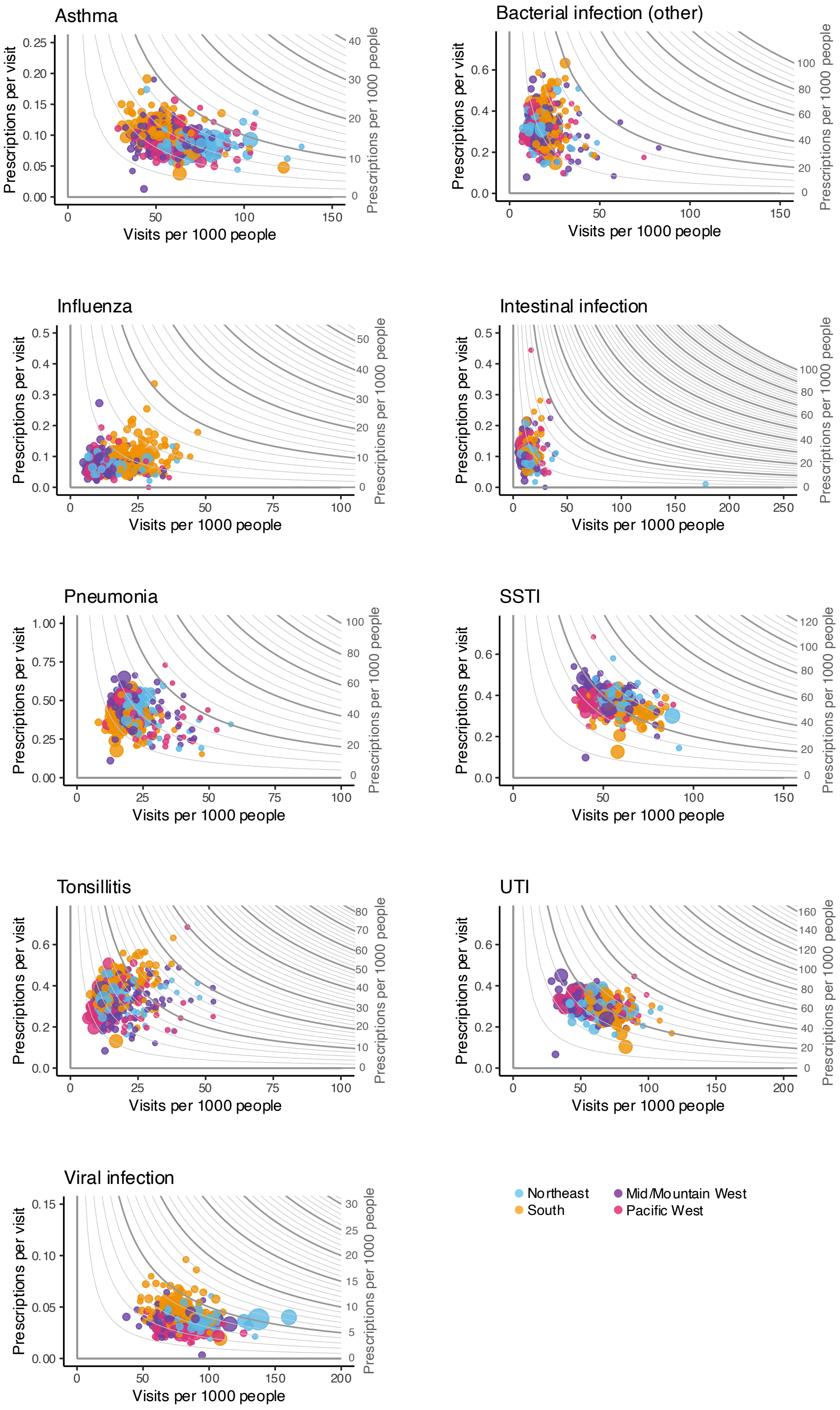

Asthma
Bacterial infection (other)
Influenza
Intestinal infection
Pneumonia
SSTI
Tonsillitis
UTI
Viral infection
Prescriptions per visit
Visits per 1000 people
Prescriptions per 1000 people
Northeast
South
Mid/Mountain West
Pacific West

**Supplementary Figure 2. Fraction of the variation in *per capita* antibiotic prescribing explained by per-visit prescribing and visits.** Lines depict the least-squares linear regression between per-visit prescriptions (blue, panel A) and outpatient visits (red, panel B) against the $log_{10}$-transformed volume of antibiotic prescriptions per 1000 people associated with each condition.

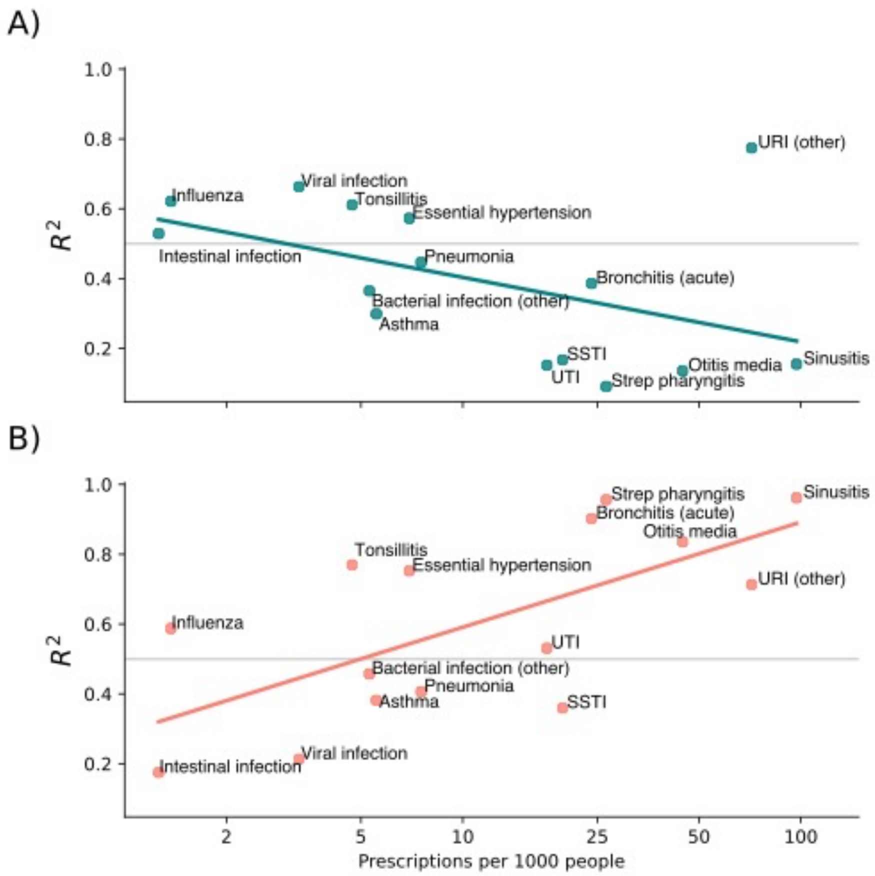
